# Dynamics of amygdala connectivity in bipolar disorders: A longitudinal study across mood states

**DOI:** 10.1101/2021.03.30.21254608

**Authors:** Gwladys Rey, Thomas A.W. Bolton, Julian Gaviria, Camille Piguet, Maria Giulia Preti, Sophie Favre, Jean-Michel Aubry, Dimitri Van De Ville, Patrik Vuilleumier

**Affiliations:** Laboratory for Behavioral Neurology and Imaging of Cognition, Department of Fundamental Neurosciences, University of Geneva, Geneva, Switzerland; Institute of Bioengineering, École Polytechnique Fédérale de Lausanne (EPFL), Lausanne, Switzerland; Department of Radiology and Medical Informatics, University of Geneva, Geneva, Switzerland; Department of Mental Health and Psychiatry, University Hospitals of Geneva, Geneva, Switzerland

## Abstract

Alterations in activity and connectivity of brain circuits implicated in emotion processing and emotion regulation have been observed during resting-state for different clinical phases of bipolar disorders (BD), but longitudinal investigations across different mood states in the same patients are still rare. Furthermore, measuring dynamics of functional connectivity patterns offers a powerful method to explore changes in the brain’s intrinsic functional organization across mood states. We used a novel co-activation pattern (CAP) analysis to explore the dynamics of amygdala connectivity at rest in a cohort of 20 BD patients prospectively followed-up and scanned across distinct mood states: euthymia (20 patients; 39 sessions), depression (12 patients; 18 sessions), or mania/hypomania (14 patients; 18 sessions). We compared them to 41 healthy controls scanned once or twice (55 sessions). We characterized temporal aspects of dynamic fluctuations in amygdala connectivity over the whole brain as a function of current mood. We identified 6 distinct networks describing amygdala connectivity, among which an interoceptive-sensorimotor CAP exhibited more frequent occurrences during hypomania compared to other mood states, and predicted more severe symptoms of irritability and motor agitation. In contrast, a limbic CAP comprising the hippocampus and ventral tegmental area exhibited fewer occurrences during both depression and hypomania compared to euthymia. Our results reveal distinctive interactions between amygdala and distributed brain networks in different mood states, and foster research on interoception systems especially during the manic phase. Our study also demonstrates the benefits of assessing brain dynamics in BD.

## Introduction

To achieve good monitoring of bipolar disorder (BD) patients’ state and prognosis, psychiatric research needs to better characterize the neural processes that distinguish different clinical phases of the disease. Resting-state brain imaging studies of BD, initially limited to Independent Component Analysis (ICA) and seed-based functional connectivity (FC), and later enriched by graph theory methods, have contributed to the demonstration of widespread FC disruptions [1–3]. This literature incriminates different brain areas and networks such as the default-mode network, limbic and reward circuits, and more recently sensorimotor networks [4–7]. In particular, the amygdala, already known for its central role in models of bipolar disorders [8,9], undergoes abnormal interactions with the medial prefrontal cortex, posterior cingulate cortex, and both ventro- and dorso-lateral prefrontal cortices at rest [10–12]. However, these findings still lack consistency and reproducibility, partly because different studies recruited patients in different clinical phases and did not perform systematic comparisons between mood states, thus hindering the distinction between trait effects (diagnosis-specific) and state effects (mood-specific). Moreover, although a few investigations segregated different states, they compared different patients [2,7,13] rather than the same patients across different phases.

Therefore, the first aim of our study was to investigate FC at rest in BD patients by obtaining repetitive/periodic scans within the *same* individuals at different phases of the disease. This approach has so far only been adopted in a couple of task-based studies [e.g. 14,15], and in one resting-state study including 10 patients scanned in both mania and euthymia [16]. However, to our knowledge, the need to perform longitudinal comparisons in the same patients across all possible states (from low through normal to high mood) is still unmet.

A second aim of our study was to adopt a novel dynamic approach by considering the “non-stationarity” of the resting brain. Since spontaneous brain FC may dynamically change over time [17,18], a number of methodological developments have tried to track these temporal fluctuations. However, most studies employed a sliding-window approach, i.e., using successive averages of connectivity values derived from between-area correlations [19]. Previous applications of the latter method to psychotic disorders showed that brain connectivity dynamics provides more discriminant information than static measures computed over a whole scanning run, in the context of automatic classification of mental disorders [20,21]. However, the sliding-window correlation approach implies a computation of FC over discrete, consecutive segments of the data, carrying certain limitations especially with regard to the effective temporal resolution of the analysis and the need for parameter selection [see review in 19].

In the present study, we propose to use an alternative method, based on “co-activation patterns” or CAPs [22], which departs from the sliding-window approach by assuming that the most informative activity of the brain can be captured by a limited amount of data [23]. Indeed, work by Tagliazucchi and colleagues demonstrated that most of the time, the brain fluctuates near the ‘critical point’ (equilibrium), and that a point process analysis (PPA), selecting only a few relevant time points (i.e., when the signal exceeds a certain threshold), can produce findings similar to those obtained when the entire set of time points is considered. In the CAP method, the PPA is applied to a single seed, by retaining solely the brain volumes (with un-thresholded activity) at time points when the seed is particularly active [22]. A temporal clustering is then applied to the selected frames. Spatial averaging of the frames grouped together yields different co-activation patterns; i.e., sets of brain areas that intermittently co-activate or co-deactivate with the seed.

Here, we applied the CAP method using the toolbox *TbCAPs* [24] to explore the dynamics of amygdala connectivity in our longitudinally followed bipolar disorder patients. The sensitivity and potential specificity of the amygdala to affective episodes in BD [2,10,25], and more generally its major involvement in emotion appraisal and emotion regulation processes [26], motivated our choice of this structure as a seed in order to probe for differential connectivity patterns across mood fluctuations. We expected that the dynamics of amygdala connectivity would not only unveil its interactions with multiple brain networks – but also reflect the clinical state of the patients, especially when comparing affective episodes with normalized mood.

## Material and Methods

### Participants

The study protocol and the informed consent procedure received approval from the ethics committee of the Geneva University Hospitals. Table 1 provides demographic and clinical description of the patients and controls samples.

**Table 1.** Demographic and clinical description of the participants.

|  | Bipolar disorders patients (BP) |  |  | Healthy controls (HC) | BP versus HC |
| --- | --- | --- | --- | --- | --- |
|  | N = 20 |  |  | N = 40 |  |
| Age | 40.4 (10.7) | | | 38.9 (10.0) | $t(58) = 0.54$, ns |
| Gender | 15m, 5f | | | 23m, 17f | $\chi^2 = 2.74$, ns |
| Handedness R/L | 17/2, 1 ambidextrous | | | 33/5, 2 ambidextrous | $\chi^2 = 0.08$, ns |
| Education, years | 11.9 (3.7) | | | 13.2 (3.3) | $t(58)=1.36$, ns |
| Diagnosis | 13 BD-I; 4 BD-II; 3 BD-NOS |  |  |  |  |
| Age onset | 19.2 (7.4) |  |  |  |  |
| Illness duration | 21.2 (10.7) |  |  |  |  |
| Mood state / group | Euthymia(E) | Depression(D) | Hypomania(H) | Controls(C) | Mood/Group comparison <sup>(a)</sup> |
| Individuals | 20 | 12 | 14 | 41 |  |
| Sessions | 39 | 18 | 18 | 55 |  |
| YMRS | 1.37 (1.67) | 0.92 (1.09) | 11.39 (4.13) | 0.46 (0.99) | $H > E = D = C$ |
| MADRS-S | 3.99 (3.26) | 14.19 (3.36) | 2.19 (2.67) | 1.17 (1.23) | $D > E > H = C$ |

Standard deviations are indicated in brackets. *BD-I*, *BD-II*, and *BD-NOS* refers to Bipolar Disorder type I, II and Not Otherwise Specified; *YMRS* Young Mania Rating Scale; *MADRS-S* Montgomery Asberg Depression Rating Scale, self-rating version. <sup>(a)</sup> Mood and group comparisons were tested using mixed model in R.

Twenty subjects with BD were recruited from the outpatient unit “*Mood Disorder Program*” of the Geneva University Hospitals. The diagnosis of bipolar disorder was based on the DSM-IV-TR criteria and confirmed by the *Mini*-*International Neuropsychiatric Interview* [MINI; 27] administered by a trained clinician (SF, CP). Twelve patients met criteria for at least one other lifetime Axis I psychiatric disorder, and eighteen patients were medicated during the follow-up study (see Supplementary Tables S1 and S2).

Forty-one healthy controls were recruited. None of these participants had a history of neurological illness or Axis I psychiatric disorders as assessed by the MINI.

### Follow-up sessions

During the follow-up period (16 ± 5 months in average), the patients completed several experimental sessions with an average interval of 3 months (± 2) between two successive sessions. Each session comprised a systematic psychometric assessment of mood including the clinician-rated Young Mania Rating Scale [YMRS; 28,29], the self-rated Montgomery–Åsberg Depression Rating Scale [MADRS-S; 30,31], and the self-rated Internal State Scale [ISS; 32]. Based on recommended cut-off scores for these scales [33, YMRS french version 29], we classified the mood state of the patients at each session into three categories labeled “hypomania” (YMRS score ≥ 6; MADRS-S score < 10), “depression” (MADRS-S score > 12, or 10 to 12 with ISS categorization into depression; YMRS < 6), and “euthymia” (MADRS-S score < 10; YMRS score < 6).

Patients and controls completed 82 and 58 MRI sessions, respectively, among which 7 and 3 were further excluded from the analyses due to excessive motion during scanning or because subjects fell asleep. Applying the previously described psychometric criteria to our sample, we obtained data from 10 patients in the three categories of mood states, from six patients in two categories, and from four patients in one category only. Table 1 (bottom part) presents an overview of the distribution of sessions included in the analyses, as well as psychometric evaluations results. Regarding medication classes, treatment regimens did not differ significantly across mood state (Supplementary Table S2). However, we further tested for changes in dosage for each single medication class in relation to our mood categories. We considered dose equivalents for antidepressants, antiepileptic benzodiazepines and atypical antipsychotics, while keeping original doses for antiepileptic mood stabilizers and testing lithium effects separately, all of which were z-scored (see Suppl. information). Higher antidepressant doses were associated with higher MADRS scores and lower YMRS scores (t=2.02, p=0.0482 and t=-2.240, p=0.0292 respectively).

### Data acquisition and analysis

For each session, 150 functional images were acquired with a TR of 2.1 s while subjects were instructed to lie awake with eyes closed, and not think about anything in particular [34]. Image processing included standard procedures implemented in SPM8 (http://www.fil.ion.ucl.ac.uk/spm). Functional images were realigned, slice-time corrected, normalized to the standard Montreal Neurological Institute EPI template, and spatially smoothed with a 5-mm kernel. We discarded image volumes with frame-wise displacement above 0.5 mm and subjects (or sessions) with more than 50% of scrubbed frames [35]. Time series of ROIs in the white matter and cerebrospinal fluid, as well as the six affine motion parameters, were used as nuisance variables to be regressed out from the data. Data were filtered between 0.01 and 0.1 Hz. For CAPs analyses, we constructed a mask based on gray matter segmentation and a probabilistic anatomical midbrain atlas including the ventral tegmental area (VTA) and the substantia nigra (SN) [36] to avoid missing reward-related brain structures relevant for BD [6].

### CAP methodology (dynamic connectivity)

Our objective was to extract amygdala co-activation patterns by selecting time points when the activity of this region is high, and then averaging brain maps at the time points that shared a similar spatial distribution of activity. For this purpose, we used a combination of the left and right amygdala masks from the AAL Atlas [37] as a seed. For each fMRI session, we extracted and Z-scored this seed BOLD time-series, and selected the 10% time points with the highest activity. We used the K-means clustering algorithm to classify the common pool of selected whole-brain volumes from all patients and controls into different clusters, for which within-cluster differences (as quantified by spatial similarity) were smaller than across-cluster differences (see [38] for more details about clustering). With a number of clusters fixed at six, the clustering showed a satisfying reproducibility level (see Supplementary Material), and yielded a set of co-activation patterns (CAPs) representing networks with potential functional relevance and/or spatial similarity with conventional resting-state networks (see Results section). These CAPs were transformed into spatial Z-maps (normalization by the standard error), so that they quantify the degree of significance to which the CAP values deviate from zero.

For each session, we computed the fraction of the retained frames assigned to each cluster – i.e., the global occurrence rate of each CAP. We also quantified the “entry rate”, that is, the number of times that the amygdala is entering a particular state of co-activation with other brain regions, regardless of whether that state is sustained for one single or several time points. We normalized the sum of entries by the total number of retained frames for the considered session. Additionally, we measured state duration (in seconds) which features the average time for which a CAP is sustained when it appears (i.e. total number of frames assigned to the cluster, multiplied by the TR and divided by the number of entries).

Statistical analyses were performed by running a linear mixed model in R, in which the variable quantifying the CAP (occurrence rate/ entry rate/ duration) was modeled as a function of mood or group (fixed factor), while subjects were modeled as a random factor. Age and sex effects and their interactions were also tested.

Finally, in the patients, we explored possible correlations between these CAP metrics and clinical scores on the MADRS and YMRS. For that purpose, we computed one Pearson correlation value for each subject taking into account all his sessions independently of the current mood, which was possible for patients having more than 2 sessions with non-zero values. A Student t-test was used to test whether the correlation values obtained in patients were different from 0. As a second step, to gain further insights into any significant correlation between mania or depression scores and a particular CAP metric, we tested the same correlation with different subscores of the main clinical scales (MADRS or YMRS), which we computed based on the weights reported in Principal Components Analysis studies in larger samples of bipolar disorder patients [39,40].

### Conventional seed-based connectivity analysis

For comparison purposes, we computed a standard amygdala seed FC map. This method computes a Pearson correlation between the spatially averaged fMRI signal in the seed, and the signal from each brain voxel included in the mask (static FC). As inputs for this analysis, we used the same masked, preprocessed data as used for CAP analyses, and the same bilateral amygdala seed. We then tested the effect of mood and group as well as correlations with depression and mania using flexible designs in SPM12.

### Control for medication effects

We also conducted further analyses to control for potential medication effects on CAP metrics by entering original or equivalents dose when appropriate as covariates in the mixed models (see Suppl. information for more details). Medication effects were also tested in the seed-based correlation analysis using medication dose as a covariate in the flexible designs.

## Results

### Static functional connectivity

We performed a standard seed-based FC analysis of the amygdala in our longitudinally followed patients and controls. We did not find any effect of the group. In the patients, however, we found a main effect of mood state in the left superior occipital gyrus/ cuneus ([-15,-85,22], Ke=58; Zmax=4.36; p=0.0112, FWE-corrected at cluster level). The activity in this area was, on average during the whole resting-state block, more strongly correlated to the amygdala fluctuations during euthymia than during depression and hypomania. Functional connectivity was not associated with depression or mania clinical scores nor with medication dosage.

### Co-activation patterns: different network interactions of the amygdala

Our dynamic connectivity analyses identified six distinct CAPs, each interacting in a distinctive manner with the amygdala seed region (see Figure 1). CAP_1_ involved an interoceptive-sensorimotor network and showed intense coactivation bilaterally in the middle insula, Rolandic operculum, and superior temporal gyrus, extending secondarily to the anterior middle cingulate cortex (MCC) / supplementary motor area (SMA), putamen, and finally (with Z score < 2) to more posterior and anterior sectors of the insula and to the central sulcus. CAP_2_ overlapped with the default mode network (DMN), comprising the posterior cingulate cortex, medial prefrontal cortex, bilateral angular gyri, and middle temporal gyri. CAP_3_ represented a ‘visual network’, largely covering the occipital lobe in its inferior, middle, and superior parts, including the calcarine area and the lingual gyrus. CAP_4_ was considered a ‘limbic network’ as it mainly included the hippocampus and parahippocampal gyrus, the VTA in midbrain, and at a lower intensity (with Z score < 3), the temporal pole and putamen. CAP_5_ constituted a ‘striatal network’, primarily involving the putamen and caudate, and secondarily the right inferior frontal gyrus (IFG) and MCC / medial superior frontal gyrus. Finally, CAP_6_ overlapped with the ‘dorsal attention network’ and comprised the inferior and superior parietal gyri bilaterally, the post-central gyri, and the SMA / MCC.

**Figure 1.**
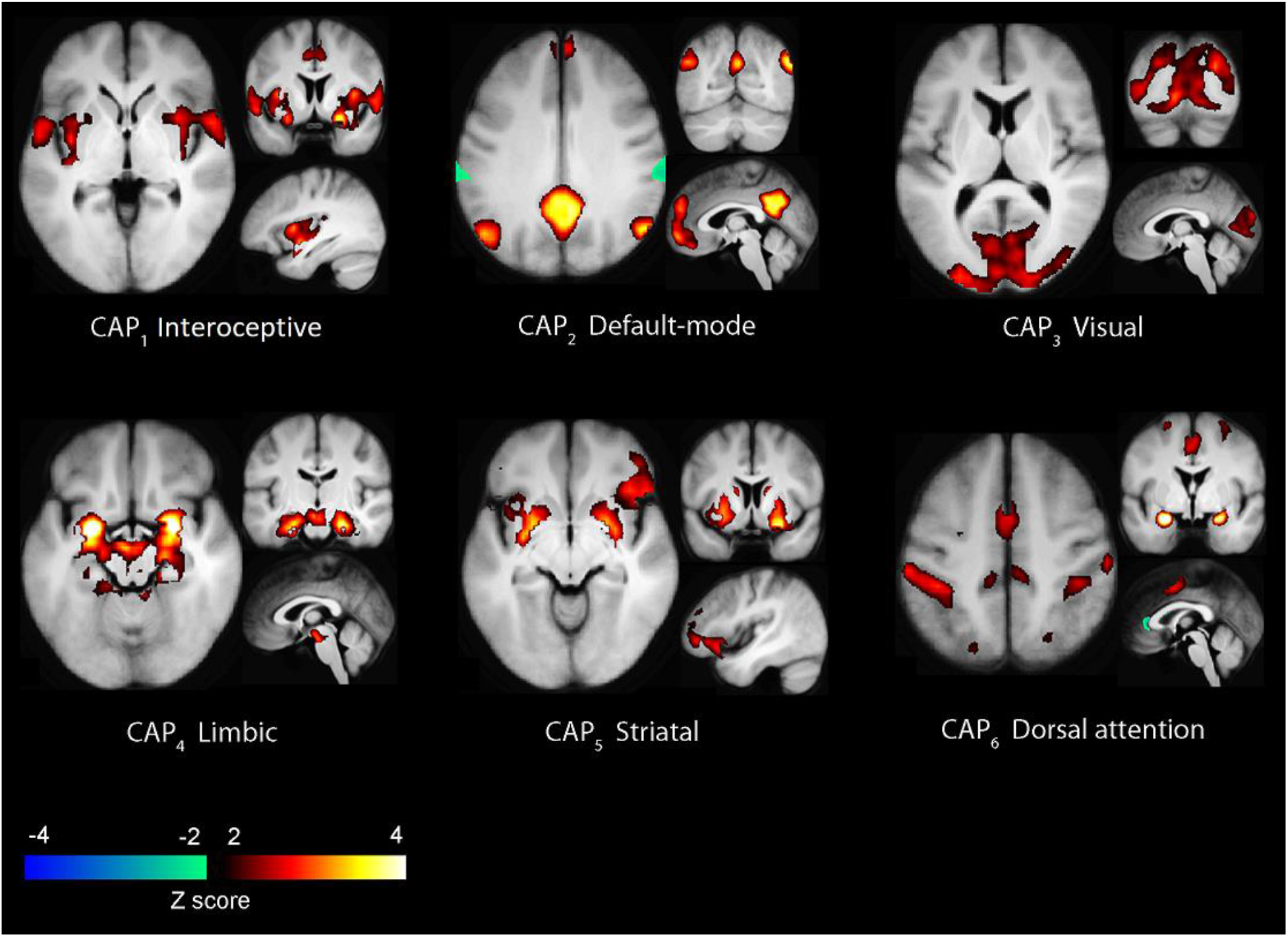
Amygdala’s co-activation patterns: from CAP_1_ to CAP_6_. Areas in hot colors co-activate while areas in cold colors co-deactivate with the bilateral amygdala seed.

### Occurrence rate, entry rate and duration of the CAPs

We then computed the temporal metrics for each CAP. We first compared the occurrence rates between each group and each mood state (Figure 2A). Any difference in the total number of time points assigned to a particular state of co-activation (occurrence rate metric) was further clarified by also testing for potential corresponding changes in the number of times the seed entered that particular state (entry rate metric) and/or by computing the mean duration of that state (duration metric). Statistically significant findings related to mood comparisons and correlations are reported in Table 2.

**Table 2.**
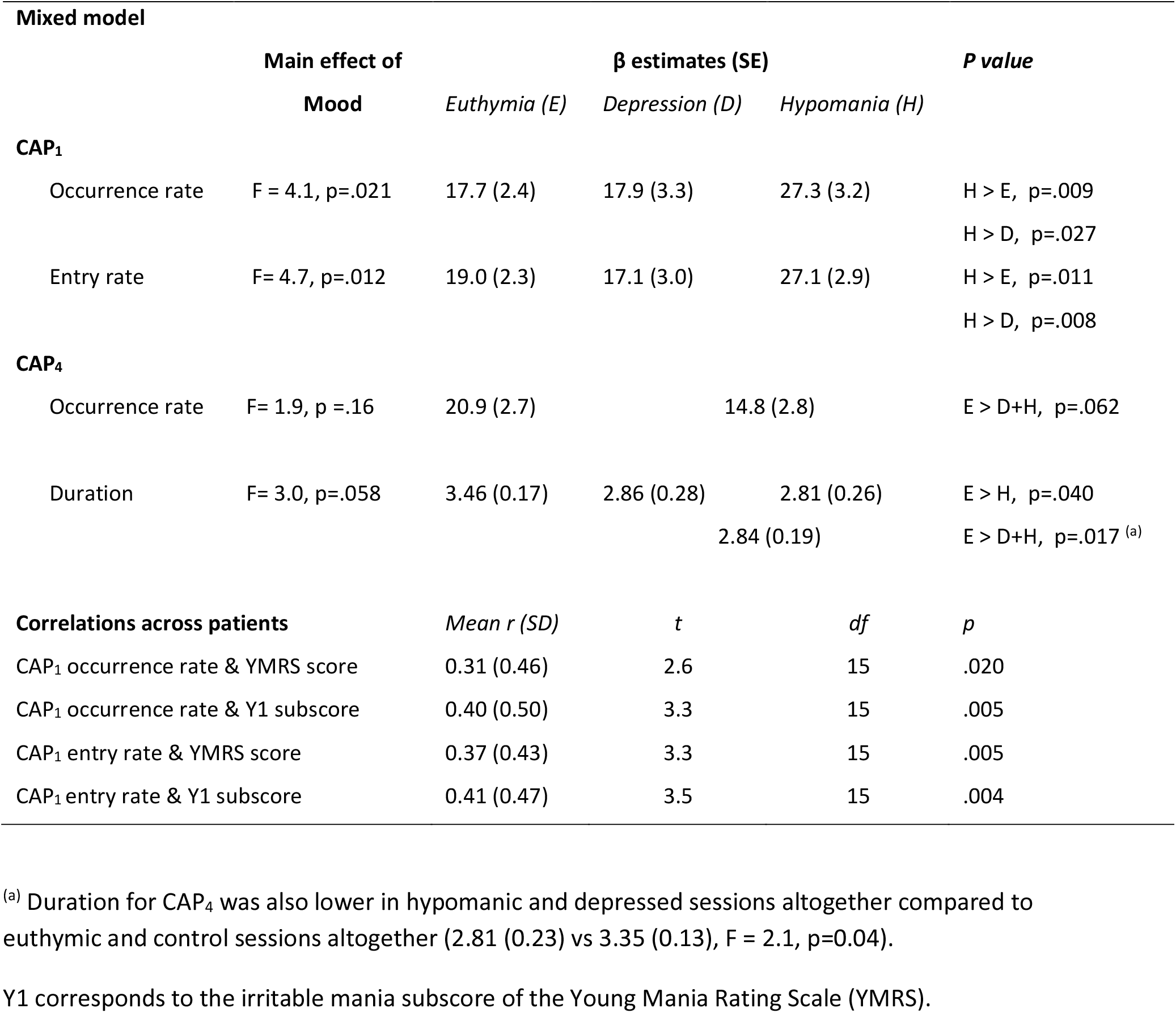
Results of the analyses of the occurrence rate, entry rate and duration of the CAPs.

**Figure 2.**
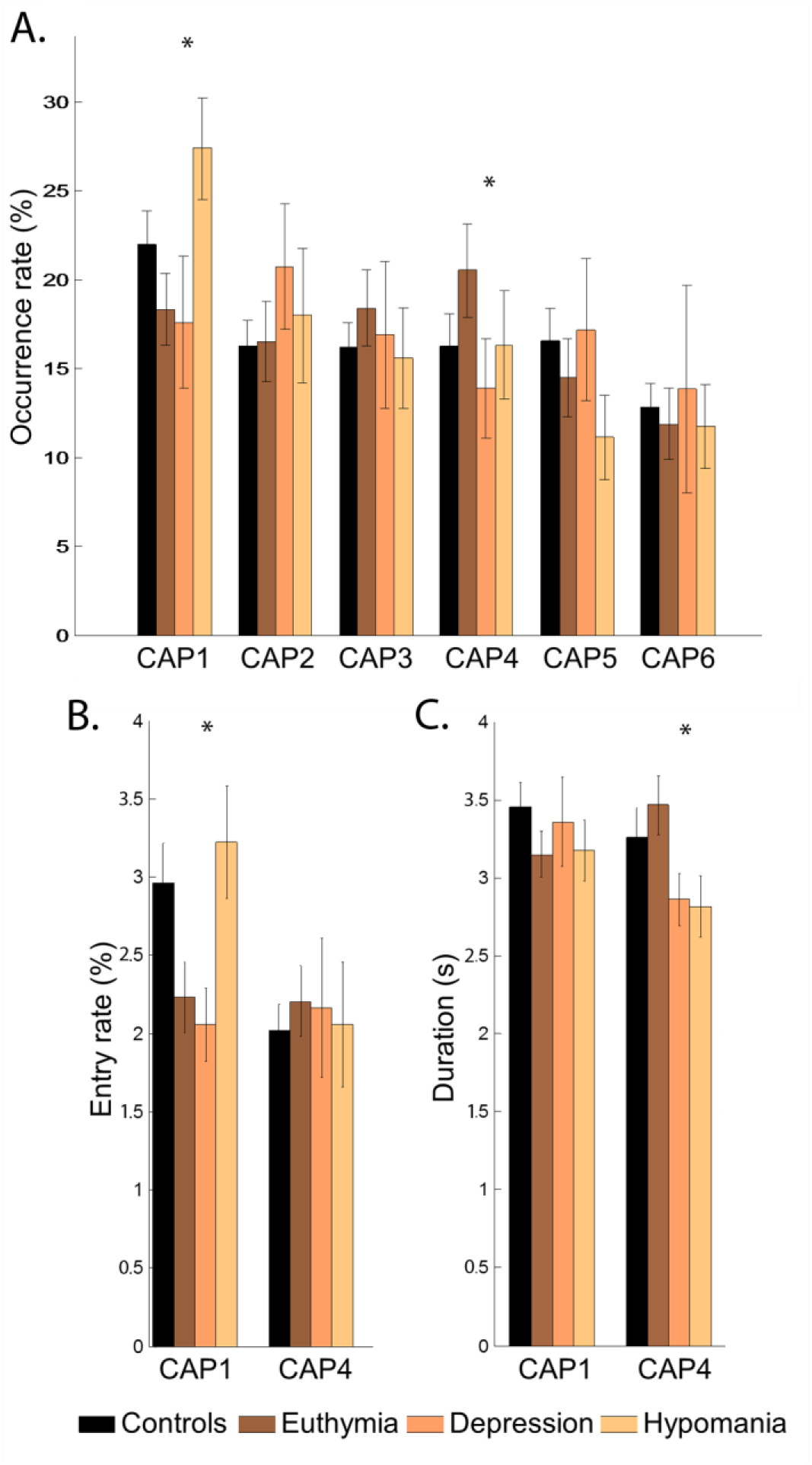
Temporal metrics of the CAPs. Occurrence rate (A), entry rate (B) and duration (C) are shown for controls and for patients depending on mood. Starts indicate significant mood comparisons within the patients group (see Results section). Error bars indicate standard errors.

For CAP_1_ (interoceptive-sensorimotor network), a mixed model analysis including the patients only revealed a significant main effect of mood on occurrence rate. Accordingly, CAP_1_ occurred more often during hypomania than during euthymia and depression. Further analyses showed that the entry rate for CAP_1_ was also increased during hypomania compared to euthymic and depressed states (Figure 2B.). In addition, across the patients, higher occurrence rate and higher entry rate for CAP_1_ were both correlated with more severe mania (total YMRS score, see Table 2). To understand the relationship between CAP_1_ and mania, we tested the same correlations by taking sub-scores of this scale representing three different aspects of mania [39]. We found that the factor ‘irritable mania’ (Y1) positively correlated with both the occurrence rate and the entry rate (Table 2), whereas ‘elated mood’ and ‘psychotic mania’ did not. By contrast, the mean duration of CAP_1_ was not affected by mood state and not related to clinical scores. Finally, comparing CAP_1_ across the 4 different participant/mood subgroups, we find a significant effect of group for the entry rate (F=2.9, p=.040), mainly driven by a higher rate in hypomanic patients than controls (ẞ (SE)=27.1(2.9) vs 20.6(1.7), p=.056). The effect of group was not significant for the global occurrence rate.

For CAP_4_ (the limbic network), despite a lack of main effect of mood, we observed an increased occurrence rate during euthymia when compared to depressed and hypomanic sessions altogether, which almost reached significance (Table 2). Furthermore, while the amygdala connectivity did not enter that state more often (no difference in entry rate), CAP_4_ lasted consistently longer during euthymia than hypomania, or depression and hypomania together (Figure 2C). The model including both patients and controls (four subgroups) did not show any significant difference in temporal metrics for CAP_4_ between controls and patients in any of the mood state.

The other CAPs did not show any effect involving group or mood on occurrence rates, and we found no effect or interaction involving age or sex.

In terms of medication, we identified a general effect of taking antidepressants, which was associated with a decrease in CAP1 occurrence rates, and a general effect of taking benzodiazepines, which was associated with an increase in CAP4 occurrences. However, doses of medication were not collinear with mood (or clinical variables) and did not interact with the effects of mood (for more information, see Suppl. information). Mood effects were still significant even after controlling for antidepressant or benzodiazepine doses effect. We did not find any effect or interaction for other medication classes on CAP1 or CAP4 metrics.

## Discussion

We explored the spatial and temporal dynamics of amygdala connectivity at rest in 20 BD patients longitudinally followed-up across different clinical phases through repetitive scanning sessions. Our analysis of static amygdala FC revealed poorly discriminating between the three categories of mood state, which is not surprising since very few data show that amygdala’s connectivity at rest is state-dependent in BD patients, be it through mood comparison studies [10,16,41] or clinical scores correlation [2]. In contrast, our co-activation pattern analysis highlighted that, among the six different networks whose activity transiently co-varied with the amygdala, two were modulated across the three clinical states during the longitudinal follow-up. First, the temporal dynamics of the interoceptive-sensorimotor CAP_1_ were consistently affected by current mood, with a higher occurrence rate of this pattern during hypomania than during the other mood states. Further analyses confirmed that the amygdala was more often engaged in functional interaction with this network, with more frequent entries into that state but no longer durations thereof. In addition, increased expression of CAP_1_ correlated with the severity of the manic symptoms. Second, the limbic CAP_4_ showed a lower global occurrence rate during affective episodes than during euthymia, in this case associated with a shorter duration of co-activation states.

CAP_1_ is predominantly composed of bilateral structures involved in interoceptive processes, namely the middle and posterior parts of the insula [42,43], which receive visceral information important for homeostatic regulation. The network also comprises elements of the sensorimotor systems including co-activated regions in the lateral and inferior sectors of somatosensory and motor cortices in the Rolandic operculum, precentral gyri, and central sulcus, as well as the putamen [44], cingulate motor areas [45], and the SMA. The more frequent co-activation of the viscerosensory areas (middle and posterior insula) and the amygdala during manic phases fuels the debate on interoception disturbances in mood disorders [46–48]. According to computational perspectives, interoception can be modeled as an iterative process of comparing the brain’s expectation of sensations with those coming from the sensory world or the internal milieu [47]. In case of discrepancy, prediction error signals are computed in the viscerosensory areas and transmitted to the visceromotor (control) areas (e.g. anterior insula and anterior ACC), which can initiate an adequate adjustment [49]. Some authors argue that mood acts as a hyperprior over emotional states, i.e., mood can bias the confidence we place in our prior beliefs relative to sensory evidence [50]. In this model, mania would be associated with hyperprecise predictions, resistant to negative feedback loops (i.e., lacking adjustment to the actual sensory signal), with prediction bias towards rewarding and predictable environment [50,51]. Given the central role of the amygdala in the predictive interoceptive circuitry [49,52], our findings are compatible with such a prediction bias, which may entail a co-occurrence of heightened arousal signals in the amygdala and increased error signals in the viscerosensory areas. Thus, the present findings bring new arguments in favor of the conceptualization of bipolar disorders as an “interoceptive psychosis” [53]. These results also converge with accumulating evidence from resting-state FC studies concerning the sensorimotor networks in BD disorders [5,13,54,see also 55] as well as increased connectivity observed between the amygdala and motor [e.g. SMA, 56] or sensory networks [among others, 2]. The dysbalancing of intrinsic brain activity toward sensorimotor patterns in mania is actually supported by prior investigations on the various phases of BD using different resting-state fMRI measurements [13,57,see also 58]. In addition, a recent model on neurotransmitters - resting-state networks interaction suggests that a concomitant over-activation of SMN and salience network (including insula) contribute to manic symptomatology [59]. Interestingly, consistent with our interpretation above, the occurrence of CAP1 in our study correlated with the factor ‘irritable mania’ from the YMRS scale – a factor that reflects, primarily, symptoms of irritability and increased motor activity / energy, and secondarily, disruptive-aggressive behavior [39]. This could result from the heightened reactivity of motivation-related error signals mediated by dynamic amygdala-interoceptive circuits. Altogether, these findings are consistent with a form of hypersalience processing [2] and perhaps denote a stronger affective meaning of somatosensory experiences and exaggerated arousal during manic states.

The second network differentially interacting with amygdala as a function of mood was CAP4, essentially composed by the anterior hippocampus, parahippocampal gyrus, and VTA. These areas constitute a limbic network encompassing parts of the mesolimbic pathway, which involves dopaminergic neurons from the VTA projecting to the nucleus accumbens, amygdala, and hippocampus, while conversely the amygdala and hippocampus both send glutamatergic innervation to the VTA. This circuit plays a major role in reward processing and addiction, but also fear conditioning and stress [60–62]. In bipolar disorders, abnormal responsiveness of the reward system has been documented, reinforcing a dopaminergic hypothesis of the disorder [63–65]. At rest, changes in FC among these structures have rarely been reported in BD [66,67]. Other studies suggested that such changes are associated with exposure to childhood maltreatment or stressful events [68–70] – known to be risk factors for mood disorders [71]. Here, by examining dynamical interactions of the amygdala in our patients, we observed that the limbic CAP_4_ tended to occur less often during depression and hypomania than during euthymia, and had a shorter duration of co-activation. This may point to some instability of this network as a marker of abnormal mood state, which could relate to impairments in motivational and reinforcement aspects of behaviour during affective episodes, normalized during euthymia (with CAP duration similar to controls). Such anomaly would also accord with previous reports of disturbances in meso/paralimbic networks in BD [2,6,67].

Two main conclusions may be drawn from this study. First, the amygdala exhibits increased transient interactions with an interoceptive-sensorimotor network during hypomania. This co-activation state appears more frequently under elevated mood states, and its association with manic symptoms seems to be driven by irritability and motor agitation. Such an association between emotion processing and interoception/sensorimotor networks might underlie the heightened arousal state of mania. Second, the amygdala also showed differential interactions with a meso-limbic network that appears less stable during affective episodes, in the sense that the connections disengage faster than during euthymia.

Several limitations should be considered, however. Although we obtained a large number of scans in all the patients through systematic prospective follow-ups, the sample size is still modest and we could not obtain analyzable scans in all the patients in each of the three categories of mood state as determined by our criteria. In addition, our exploration of medication effects on brain dynamics should be considered with caution, as it is presently limited by the heterogeneity of patients’ medication regimens, the small number of patients taking each medication class, and unassessed potential drug interactions.

Nevertheless, this study allowed us to highlight the relevance and high potential of the CAP analysis especially for investigating the dynamics of neural circuits coupled with emotional processing systems such as the amygdala, and thus unveil functional disturbances in brain systems subtending interoceptive/sensorimotor and reward/motivation processes, whose physiopathological significance is currently gaining interest in BD.

## Data Availability

Under direct request to the corresponding author

## Acknowledgements

We thank Djalel Eddine Meskaldji (EPFL) and Rémi Neveu (UNIGE) for general discussion and help with the statistics. We also thank the patients for taking part in this longitudinal study.

## Author Contributions

GR, PV, DV conceived and designed this research, and interpreted the data; GR, TAWB, MGP and JG analyzed the data; GR, CP, SF and JMA acquired the data; GR wrote the paper, and all authors reviewed the final submission.

## Additional Information

Supplementary Information accompanies this paper.

## Supplementary Information

### Methods

#### Data acquisition

Neuroimaging data were collected using a 3T Magnetom TIM Trio scanner (Siemens, Germany) and a 32-channel head-coil. The Blood Oxygenation Level-Dependent (BOLD) contrast was evaluated using a T2*-weighted echo-planar sequence (EPI). Five hundred functional volumes of 36 axial slices each (TR/TE/flip angle = 2100 ms/30 ms/80°, FOV=192 mm, resolution=64×64, isotropic voxels of 3.2 mm^3^, distance factor 20%) were acquired in one single continuous scanning run. We collected a high-resolution T1-weighted anatomical image (TR/TI/TE/flip angle=1900 ms/900 ms/2.27 ms/9°, FOV=230 mm, resolution=256×256, slice thickness=0.9 mm, 192 sagittal slices) at the end of the first session.

#### Clustering reproducibility

To estimate an optimal number of clusters into which to subdivide the fMRI volumes with strong amygdala activation, we randomly split our full subject population (controls and BD) into two equally sized groups for a total 100 times; clustering was performed each time, separately on each half, over a range of cluster numbers from 2 to 12. Best-match pairs between each group’s generated clusters were established with the Hungarian algorithm [1], and mean spatial similarity, averaged over the 100 trials, was considered as our reproducibility measure.

To assess the significance of this measure with regard to an appropriate null distribution, we created 1’000 sets of surrogate data under a stationarity assumption: white noise with the same variance as the real data was added to a subject-specific stationary activity pattern estimated from the first eigenvector of its spatial covariance matrix, and randomly scaled over time. Following temporal Z-scoring of this surrogate data, a null distribution for mean reproducibility was generated for each cluster number, from 2 to 12, as described above.

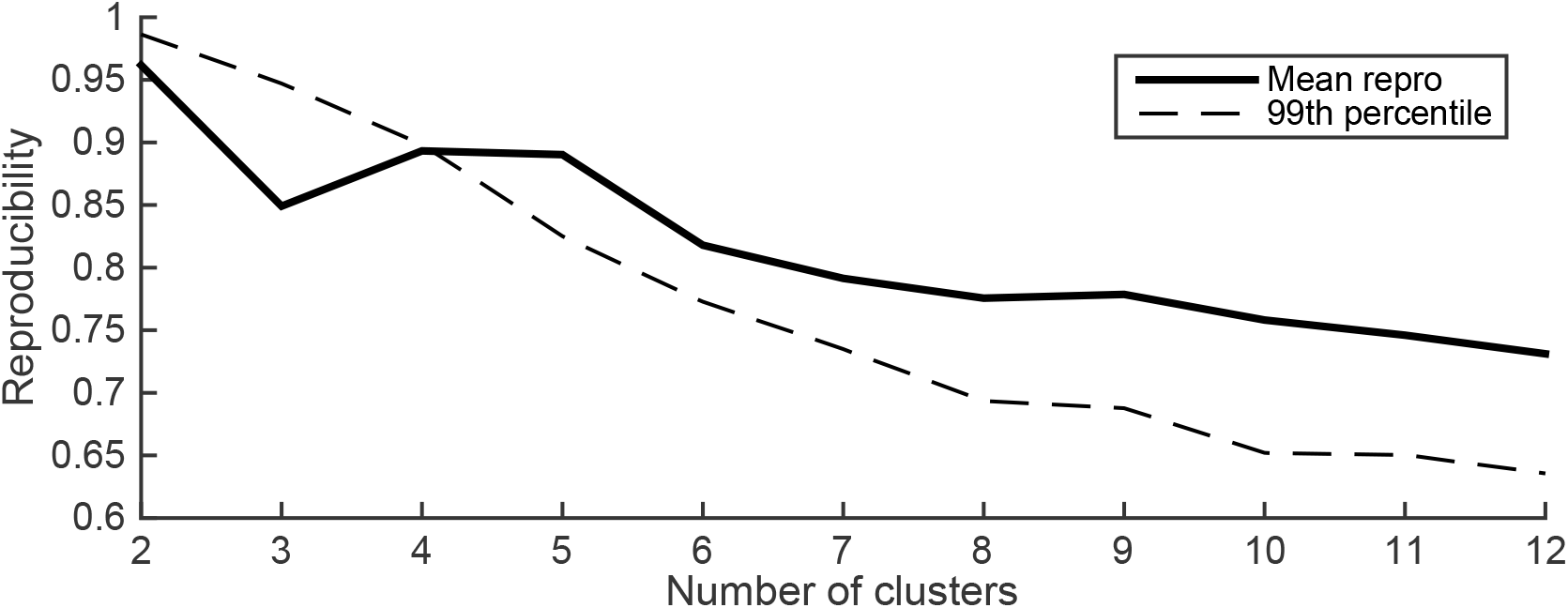

As seen in this figure, all cluster numbers larger than 4 in the assessed range of values display a mean reproducibility that exceeds the 99^th^ percentile of the associated null distribution, which means that relevant dynamic information lies in the data at all those different scales of analysis. The difference then resides in the increased subdivision of obtained co-activation patterns into smaller subnetworks. Here, we opted for a trade-off value of 6, as observed co-activation patterns matched well-known resting-state networks examined in previous functional connectivity studies.

#### Control for medication effects

To estimate dose equivalents for each medication class, we used chlorpromazine-equivalent dose for atypical antipsychotics (quetiapine fumarate, aripiprazole, olanzapine, risperidone, amisulpiride), clonazepam-equivalent dose for antiepileptic benzodiazepin (pregabalin, lorazepam, oxazepam), and escitalopram-equivalent for antidepressants (bupropion, venlafaxine, paroxetine)[2,3,4].

We kept original dosage for antiepileptic mood stabilizers (valproate and lamotrigine) and analyzed lithium effects as a single variable, all of which were z-scored before entering the mixed models (CAP metrics analysis) or flexible model (seed-based correlation analysis).

### Results

#### Effects of medication on CAP metrics

We observed a main effect of antidepressants; the five patients taking antidepressants (19 sessions) had globally fewer occurrences of CAP1 than the other 15 patients (56 sessions), with CAP1 occurrence rates at 11,6% ±11,7 (with antidepressants) against 23,3% ±13,1 (without). Considering the lack of collinearity between mood (or clinical scores) and antidepressants, and the lack of interaction between them (F=0.3, p=0.7), we applied a generalized additive mixed model which revealed a main effect of antidepressants (F=10.4, p=0.004), in addition to a main effect of effect mood (F=3.6, p=0.034). In addition, we noticed that there was no effect of antidepressant dose on occurrences in the patients who were taking this medication (F=0.4, p=0.5). For benzodiazepines, we found a main effect on CAP4 metrics, with greater occurrences of CAP4 during the 15 sessions (4 patients) with benzodiazepine than during the other 60 sessions (17 patients) (29,1% ±15,4 versus 15,1% ±13,3 respectively). Using the same methodology, an additive model was fitted, yielding a main effect of benzodiazepines (F=11.2, p=0.004) additionally to our remaining general effect of mood stability (euthymics vs depressed and hypomanics together) (F=4.58, p=0.036). Thus, differences between mood states on CAP1 and CAP4 occurrences were still significant when we controlled for medication effects by adding covariates in the models.

---

**Table S1.** Detailed clinical description of the patients. Twelve patients met criteria for at least one other lifetime Axis I psychiatric disorder.

| ***Lifetime presence of*** | ***Number of patients*** |
| --- | --- |
| ***- Axis 1 disorder:*** |  |
| General anxiety disorder | 4 |
| Panic disorder | 1 |
| Post-traumatic syndrome disorder | 1 |
| Obsessive-compulsive disorder | 1 |
| Social phobia | 2 |
| Attentional deficit and hyperactivity disorder | 3 |
| Substance use disorder<sup>1</sup> | 10 |
| ***- Other characteristics*** |  |
| Psychotic symptoms | ≥8 |
| Suicide attempt(s) | ≥5 |

<sup>1</sup> including alcohol (1 past, 3 current), cannabis (2 current, 1 stopped during the study), cocaine (2 past), ecstasy (1 past), opiate (1 past), benzodiazepine (1 past). The patients were asked to come sober the day of the experiment (sometimes short-delay appointment).

**Table S2.** Table of contingencies depicting the number of sessions during which patients were taking medication, as a function of mood and pharmacological class.

| Mood state<br>(n sessions) | Mood<br>stabilizer |  | Anti-<br>psychotic |  | Anti-<br>depressant |  | Benzo-<br>diazepine |  | Psycho-<br>stimulant |  |
| --- | --- | --- | --- | --- | --- | --- | --- | --- | --- | --- |
|  | *on* | *off* | *on* | *off* | *on* | *off* | *on* | *off* | *on* | *off* |
| Depression (n=18) | 14 | 4 | 11 | 7 | 7 | 11 | 2 | 16 | 2 | 16 |
| Euthymia (n=39) | 26 | 13 | 24 | 15 | 8 | 31 | 7 | 32 | 5 | 34 |
| Hypomania (n=18) | 14 | 4 | 12 | 6 | 4 | 14 | 6 | 12 | 3 | 15 |

Pharmacological treatment included mood stabilizers (lithium, valproate, lamotrigine), antipsychotics (quetiapine, fumarate, aripiprazole, olanzapine, risperidone, amisulpiride), antidepressants (escitalopram, bupropion, venlafaxine, paroxetine), anxiolytics (clonazepam, lorazepam, oxazepam, prégabaline), psychostimulant (methylphenidate). Treatment regimens did not differ between euthymia, depression and hypomania ( $\chi^2 = 7.7$ , $df = 18$ , $p\text{-value} = 0.98$ ).

